# Prevalence of catheter associated Urinary Tract Infection (UTI) in hospitalized patient in Karachi

**DOI:** 10.1101/2024.06.30.24309719

**Authors:** Syed Rohan Ali, Moosa Abdur Raqib, Shahida Kashif, Muhammad Ashir Shafique, Abdul Haseeb, Aisha Anis

**Affiliations:** Liaquat College of Medicine and Dentistry; Jinnah Sindh Medical University; Liaquat College of Medicine and Dentistry, Karachi; Jinnah Sindh Medical University, Karachi

## Abstract

**Background:** Catheter-associated urinary tract infections (CAUTIs) are a prevalent healthcare-associated infection, accounting for significant morbidity, mortality, and increased healthcare costs.

**Method:** This is a cross-sectional study of patients diagnosed with UTI associated with catheter use. The sample was collected from November 2023 to June 2024, consisting of 200 patients admitted to the surgical, medical, and trauma wards of tertiary hospitals in Karachi, namely Jinnah Postgraduate Medical Centre Karachi and Dr. Ruth K. M. Pfau Civil Hospital Karachi. Data is analyzed using SPSS Version 22 and P-value of 0.05 considered significant.

**Result:** The majority of respondents (59.5%) had their catheters changed since insertion, predominantly by trained nurses (93.0%). There were notable associations with underlying conditions such as hypertension (56.5%) and diabetes (44.5%). Gender differences were significant, with females leading in medical cases and males in surgical and trauma cases (p-value 0.017). Age-related trends showed the 55+ age group dominated medical cases, while surgical and trauma cases varied by age group. There was a significant relationship between bleeding during catheterization and UTI (p-value: 0.000).

**Conclusion:** The study revealed a minimal incidence of CAUTI in Karachi’s tertiary care hospitals, indicating effective practices. However, further research is needed to explore the potential risk factors identified, such as female gender and comorbidities, to develop targeted interventions for reducing CAUTI incidence and improving patient outcomes.

## INTRODUCTION

Urinary tract infections, commonly known as UTIs, are a prevalent type of bacterial infection, accounting for an estimated 25-40% of all nosocomial infections. It is reported that 70-80% of these infections are a result of using an indwelling urethral catheter.^1^

Catheter-related urinary tract infections (CAUTIs) are a type of urinary tract infection that affects individuals who have a catheter or have had one within the past 48 h. These infections are the most prevalent type of healthcare-associated infections.^2^

Risk factors for catheter-associated urinary tract infections (CAUTIs) comprise age, gender (female), diabetes, and extended catheterization duration.^3^ The most critical aspect in the occurrence of bacteriuria is the duration of catheterization, which poses a daily risk of 3– 7%.(4) ICU patients with CAUTIs experience longer hospital stays, higher healthcare costs, and excessive antibiotic use.^5,6^

Urinary tract infections (UTIs) can result from Gram-negative and Gram-positive bacteria, along with fungi. Among these, Uropathogenic Escherichia coli (UPEC) is the predominant pathogen responsible for both non-complicated (75%) and complicated (65%) UTIs.(7) Complicated UTIs, mainly caused by CAUTIs, involve common pathogens like Enterococcus spp., Klebsiella pneumoniae, Candida spp., and others, with antibiotics being the cornerstone of treatment despite biofilm-related resistance and potential microbiota alterations.^8^

UTI signs and symptoms can encompass heightened bladder awareness, urgency, frequent urination, discomfort during urination, urinary tract pain, tenderness in the lower abdomen, fever, chills, changes in mental alertness, overall fatigue, unexplained lethargy, back or side pain, and tenderness around the ribs and spine junction.^6,9,10^

A cross-sectional study conducted at the Department of Microbiology, Abbas Institute of Medical Sciences, Muzaffarabad, from January to December 2019 revealed that 143 out of 270 patients (52.96%) developed catheter-associated urinary tract infections as a result of prolonged catheterization lasting more than 48 hours.^11^

The objective of our study was to establish the prevalence of CAUTI in Karachi and to identify the most prominent risk factors in our region.

## METHODOLOGY

A cross-sectional study conducted on a sample size of 200 patients catheterized for 2 or more days. The sample was taken through nonprobability purposive sampling from Dr. Ruth K. M. Pfau Civil Hospital Karachi, Jinnah Postgraduate Medical Center Karachi, Sindh Employees Social Security Institution (SESSI) within a period of 7 months from November 2023 to June 2024. Informed verbal consent was obtained from the patient. Pilot study was conducted to assess the validity of the questionnaire. Data was analyzed using SPSS Version 22 with a 95% confidence interval, the margin of error is taken as 5% and a P-value of 0.05 was considered significant.

## RESULT

The frequency data provide a comprehensive overview of various aspects related to catheterisation and associated conditions. It was observed that the majority of respondents had their catheters changed since insertion (59.5%), and the procedure was often performed by trained nurses (93.0%). Additionally, while a significant proportion reported foul-smelling urine after catheterisation (37.5%), fewer experienced fever or chills (36.5%) or a burning sensation during micturition (51.0%). Furthermore, a considerable number of respondents had a history of urinary incontinence before catheterisation (40.5%), while 45.0% reported a previous urinary tract infection (UTI). Moreover, the data revealed associations with underlying health conditions, such as hypertension (56.5%) and diabetes (44.5%). Additionally, a notable proportion of respondents had undergone surgical procedures involving urinary catheterization for two or more days (51.5%).

A comprehensive overview of catheterisation cases categorised by age group and reasons for the procedure was presented. Notably, the 55+ age group dominated medical cases (39.39%), while surgical cases peaked in the 46-55 age group (24.24%), and trauma cases exhibited variability, with the 26-35 age group leading (31.43%). At a p-value of 0.003. Gender-wise distribution also showed notable differences, with females leading in medical cases (74), while males dominated surgical (20) and trauma (24) cases. A p-value of 0.017 indicated significant sex-related disparities in catheterisation reasons.

Moreover, the analysis of bleeding during catheterisation, categorised by UTI, revealed a significant relationship (p-value: 0.000) between bleeding and UTI. Further analyses of various factors, such as catheter change, foul-smelling urine, fever or chills, burning sensation during urination, urine incontinence, history of UTIs, hypertension, diabetes, medication changes post-antibiotics, and white blood cell count in urine samples, all showed statistically significant associations with UTI. These associations are supported by p-values of 0.001, 0.000, 0.000, 0.000, 0.000, 0.000, 0.005, 0.001, and 0.000, respectively, indicating significant relationships in each case.

## DISCUSSION

Healthcare-associated infections (HAIs), which are caused by various factors such as inadequate hospital facilities and the use of invasive procedures such as catheters and probes, pose a significant global threat. One of the most prevalent HAIs is Catheter Associated Urinary Tract Infection (CAUTI), which can lead to increased mortality and morbidity, extended hospital stays with increased costs.^12,13^ Indwelling urinary catheters and extraneous equipment are the leading causes of more than 70% of urinary tract infections, particularly when they are inserted unnecessarily or left in the bladder for an extended period.^14^

This cross-sectional study examined the prevalence and risk factors of catheter-associated urinary tract infections (UTIs). Our findings indicate that the frequency of UTIs in approximately 200 patients was 27% and that current practices in Karachi’s tertiary care hospitals are up-to-date and effective.

In a study conducted at the Department of Microbiology, Abbas Institute of Medical Sciences, Muzaffarabad, from January to December 2019 revealed that 143 out of 270 patients (52.96%) developed catheter-associated urinary tract infections as a result of prolonged catheterization lasting more than 48 hours.^11^

In another cross sectional study of about 179 patients conducted in department of medicine Muhammad Teaching hospital Peshawar showed CAUTI according to their culture reports.E.coli in 37 %,staphlyoccous epiderdimis 18 %,Pseudomonas aeruginosa 16%,Klebsellia pneumonia 20 %,proteus mirabilis 3% and Enterocoocus 6%.^15^

This above findings were similar to another study done in Peshawar at Two tertiary care hospitals showing that gram negative organism account for 79 % mainly E.coli and Enterococcus in gram positive about 21%.^16^

Our study was conducted at government hospitals in Karachi, which is why a routine urine culture report was not conducted as part of the investigation. However, most patients are diagnosed and treated with empirical antibiotics.

Our study revealed that females were more likely to experience CAUTI than males, a finding that aligns with research conducted at a Tertiary Care Hospital in Uttarakhand, India. This study involved a total of 468 patients and revealed that the prevalence of CAUTI was higher in females (53%) compared to males (47%).^17^ This could be a risk factor for CAUTI whichrequirese further research.

Our research uncovered that approximately 56.5% of the patients had hypertension and around 44.5% had diabetes, which may have a connection to CAUTI, as demonstrated in a comparable study where hypertension was found to be around 32% and diabetes was approximately 21%.^18^

Our findings revealed a minimal incidence of CAUTI in tertiary care hospitals in Karachi, demonstrating that patients are receiving adequate care in these institutions. To gain a more comprehensive understanding of CAUTI and prevent its occurrence, we suggest that additional research be conducted to explore the risk factors identified in this cross-sectional study. To further elucidate the complexities of CAUTI and develop effective strategies for its prevention, it is crucial to conduct longitudinal studies that can provide a more in-depth analysis of the identified risk factors.

## Data Availability

will be granted access by the principle investigator

## Declarations

### Funding

The authors received no extramural funding for the study.

### Author Contribution

Concept & Design of Study : Syed Rohan Ali

Drafting: Moosa Abdur Raqib

Data Analysis: Syed Rohan Ali

Revisiting Critically: Shahida Kashif, Muhammad Ashir Shafique and Abdul Haseeb

Final Approval of version: Muhammad Ashir Shafique and Abdul Haseeb

Moosa Abdur Raqib, Syed Rohan Ali and Muhammad Ashir Shafique played crucial roles in refining the content, conducting a final round of editing, and presenting the article in a structured and balanced format, ensuring it was ready for publication.

IRB Reference no : IRB-

Ethics approval granted by IRB Committee Liaquat College of Medicine and Dentistry, KHI Verbal consent was obtained from the participates.

Data was collected by the investigator. If required data and materials access can be granted.

Conflict of Interest: The study has no conflict ofinterest to declare by any author.

## Tables

**Table 01:** DISTRIBUTION OF PATIENTS.

| Reason of catheterization | Percentage % |
| --- | --- |
| Medical(132) | 66.0% |
| Surgical (33) | 16.5% |
| Trauma(35) | 17.5% |
| **Gender** |  |
| Male (102) | 51.0 % |
| Female(98) | 49.0% |
| **Age** |  |
| 18-25 (27) | 13.5% |
| 26-35 (30) | 15.0% |
| 36-45 (38) | 19.0% |
| 46-55 (37) | 18.5% |
| 55+ (68) | 34.0% |
| **Duration of catheter** |  |
| 2-4 days (37) | 18.5% |
| 4-6 days (90) | 45.0% |
| >6 days (73) | 36.5% |

**Table 02:** PERCENTAGE OF CAUTI.

|  |  |
| --- | --- |
| CAUTI ( 57) | 28.5 % |
| No CAUTI (143) | 71.5% |

## Notes

### Competing Interest Statement

The authors have declared no competing interest.

### Funding Statement

no funding recieved

### Author Declarations

Ethics approval granted by IRB Committee Liaquat College of Medicine and Dentistry, KHI

